# Applications of Behavioral Economics to HIV Programs in Low and Middle Income Countries: A Scoping Review Using the EAST Framework

**DOI:** 10.1101/2023.11.02.23298003

**Authors:** Elizabeth Long, Jacqueline Devine

**Affiliations:** DTA Innovation; USAID

## Abstract

Despite significant gains and successes in many areas, progress in HIV prevention and treatment is uneven, highlighted in the 2022 UNAIDS report *In Danger*, and certain populations are being left behind. In particular, men and adolescent girls and young women (AGYW) are among the groups in danger of not reaching epidemic control targets. Among other calls for utilizing scientific innovations, PEPFAR’s Office of the Global AIDS Coordinator and Health Diplomacy recently highlighted the need to expand the use of innovative methods from behavioral science in HIV programs. One of these innovative, but underused, approaches is behavioral economics (BE), which leverages our predictable cognitive biases and mental shortcuts to both diagnose behavioral factors and positively influence behavior. The tools used by BE to change behavior are frequently called nudges, which tend to be low-cost and easy-to-implement interventions. This scoping review identified nudges applied to select client and provider behaviors along the HIV prevention, testing, and care and treatment continuum. It maps them to the commonly used EAST Framework, a simplified way to classify nudges for program design and highlights those that shifted behavior. The EAST Framework groups nudges into Easy, Attractive, Social, and Timely categories. This scoping review identified that Easy nudges were commonly applied as changes to the structure of HIV programs, influencing individual behavior through program changes. Nudges that directly targeted individuals most commonly fell into the Attractive and Social categories. Many of the individual-focused nudges in the Attractive and Social categories changed behavior, along with Easy nudges. Additionally, the mapping highlighted the dearth of nudges applied to provider behavior in the Low and Middle Income (LMIC) HIV space. Further integration of nudges into HIV programs and their evaluation using implementation science may help move the needle to end the HIV epidemic as a public health threat by 2030.

## INTRODUCTION

The 2022 UNAIDS report In Danger highlighted the fact that progress in HIV prevention and treatment is uneven, despite monumental successes and gains in many areas.^1^ The COVID-19 epidemic has further strained resources for global health and epidemic control in HIV and there is even emerging evidence that infection rates are increasing after many years of decreasing.^1^ Certain populations, especially men and adolescent girls and young women (AGYW), are among the groups with uneven progress contributing to countries being at risk of not meeting 95-95-95 targets by 2030. In Sub-Saharan Africa, AGYW are three times more likely to become infected than adolescent boys and young men of the same age group.^1^ Around the world, only 70% of men living with HIV are receiving antiretroviral treatment compared to 80% of women.^1^

The field of behavioral science has a long and storied history with HIV programs. More traditional approaches such as Information, Education, and Communication (IEC) failed to shift behavior early in the epidemic. However, approaches such as human-centered design (HCD) and social marketing have increasingly been applied and have positively changed behavior. But within the field of Behavioral Science, the innovative approaches and tools of behavioral economics (BE) have been, to date, surprisingly underutilized.

At the core of behavioral economics is Dual Process Theory, also called System 1 and System 2 thinking, which won Daniel Kahneman the Nobel Prize in Economics in 2002.^2^ (The theory was jointly developed by Daniel Kahneman and Amos Tversky, but Nobel Prizes are not awarded posthumously therefore Amos Tversky did not win it.) The theory posits that people have two distinct ways of thinking and making decisions: System 1 is quick, instinctual, and automatic while System 2 is slow, deliberate, and more calculating.^2^ Neither System 1 nor System is ‘better’ than the other; they both serve people but in different contexts and in different ways. During any given day humans switch between Systems 1 and 2 frequently throughout the day. However, when people can or do not switch between them predictable and systematic errors in thinking emerge. These errors are called behavioral and cognitive biases and result in humans being ‘predictably irrational’.^3^

Applied behavioral economics has two key elements. The first aim is to diagnose when the predictable biases are negatively influencing people’s behaviors and the second aim is to use a set of tools and techniques to address the biases and positively influence behavior. The tools and techniques to influence behavior are frequently called ‘nudges’.

Countless nudges are being applied to behaviors around the world and a simplified way to categorize nudges is by using the EAST Framework, developed in 2014 by the Behavioral Insights Team of the UK Government.^4^ EAST stands for Easy, Attractive, Social, and Timely. In addition to helping categorize the types of nudges that have been applied in HIV programs, the EAST Framework can be a guide for how nudges can be applied to ongoing or future programs.

The scope of this article is twofold: (i) to inform HIV policy makers and program managers of the range of nudges that have been applied to specific behaviors along the prevention to treatment continuum and (ii) to highlight which nudges were effective and provide depth on a handful of interventions.

This scoping review adds to the literature by focusing on HIV behaviors and populations in LMICs for which progress has been uneven. Focusing on these behaviors reduces the number of included studies, but the narrower lens allows for a more detailed and nuanced discussion of the nudges and their effects in areas where policy makers and program managers are actively seeking innovative solutions. This review is also the first scoping review of nudges and HIV programs to detail the findings of the research and not simply categorize the types of nudges that have been tested or implemented.

## METHODS

### Search Strategy

This scoping review focused on literature related to behavioral economic tools (i.e., nudges) and their application to HIV programs. Google Scholar and PubMed were used as the search engines. Keywords of HIV plus common behavioral science and nudge terms were used to identify literature, for example, “HIV and nudge”, “HIV and behavioral economics”, “HIV and present bias”, or “HIV and social proof.” The full list of keywords is included as Supplement I.

### Inclusion Criteria

No time bounds were put on literature; however, since BE is a nascent field, the majority of publications were from the past decade. Only publications in English were included. All study types were included, including qualitative studies and reviews.

### Study Selection

EL then screened the article titles and abstracts for inclusion. The literature needed to include one of the following target behaviors and populations along the HIV continuum: increasing uptake of Pre-Exposure Prophylaxis (PrEP) among adolescent girls and young women (AGYW); increasing uptake of testing services for HIV among men; increasing initiating treatment immediately following a positive test (i.e., linkage) among men and other populations; increasing re-engagement in treatment; reducing interruptions in treatment among adolescents and young adults, young men and women, older adults, or older adults; or improving people-centric and respectful behavior by clinic-based and community-based healthcare workers. These target behaviors were identified because these populations are not reaching targets as quickly as expected, as was highlighted in UNAID’s report In Danger.^1^ Literature on populations living in Low and Middle Income Countries according to World Bank classification was included. Literature that was adjacent to the focus target behaviors and populations was also reviewed.

Following the initial search, snowball referencing was conducted by EL. References from identified studies and articles which cited them were also reviewed for inclusion.

To provide a comprehensive assessment of applications, nudges applications that were not tested and reported in published studies, such as Universal Test and Treat, are also categorized and discussed.

The search resulted in 1,174 total articles, of which 34 were included in this scoping review (Figure 1). The most common reasons for exclusion included that the tested nudges focused solely on financial incentives or non-monetary rewards or that the study did not include a relevant target behavior or population.

**Figure 1:**
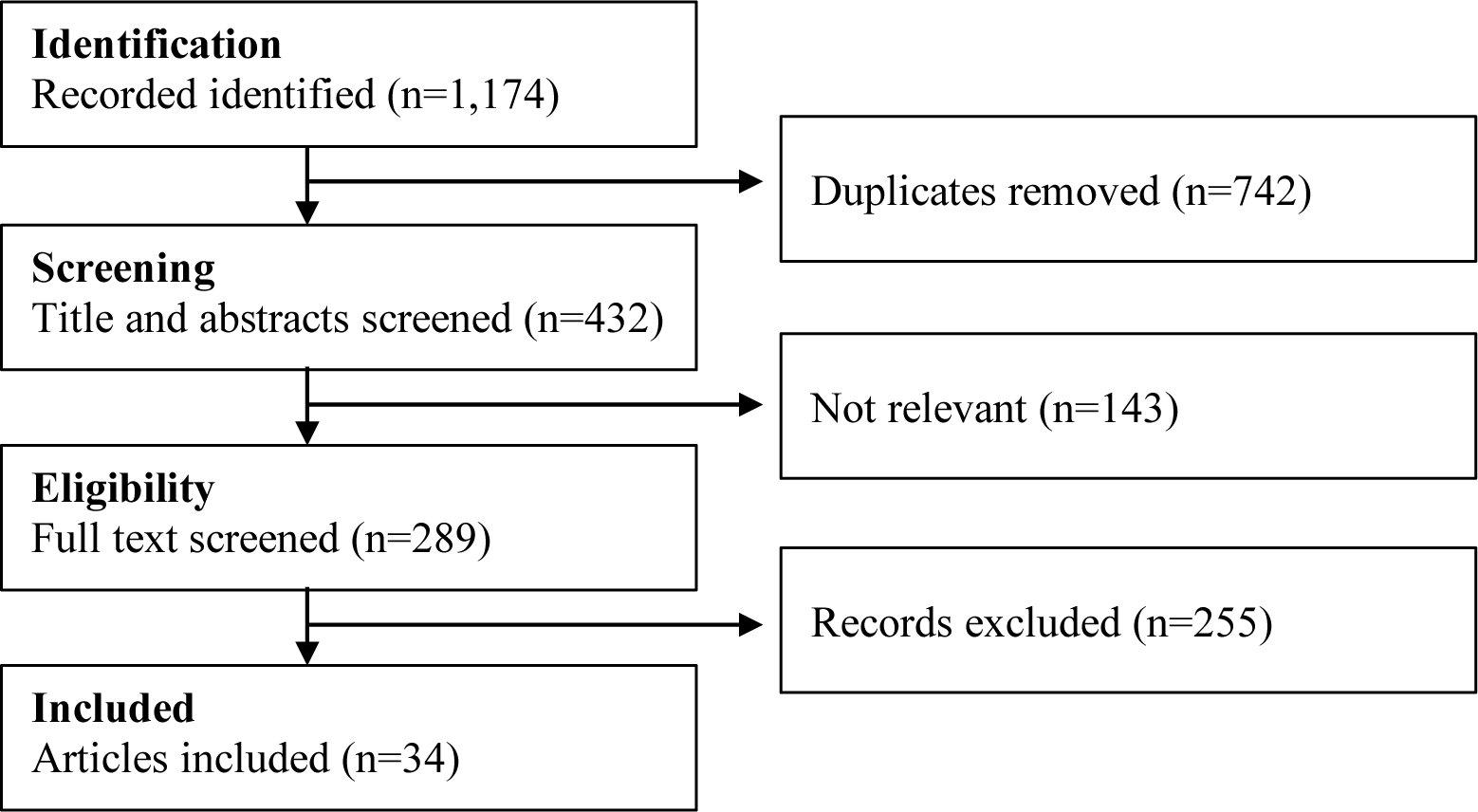
PRISMA Flowchart of Study Selection Process.

## RESULTS

Of the 34 articles, 16 described studies in East Africa while 9 described studies in Southern Africa. Only three studies occurred in LMICs outside of Africa (Table 1). In terms of behaviors along the HIV continuum, most studies dealt with either increasing continuity and re-engagement in treatment (41%) or increasing uptake of testing services (38%) (Table 1).

**Table 1:**
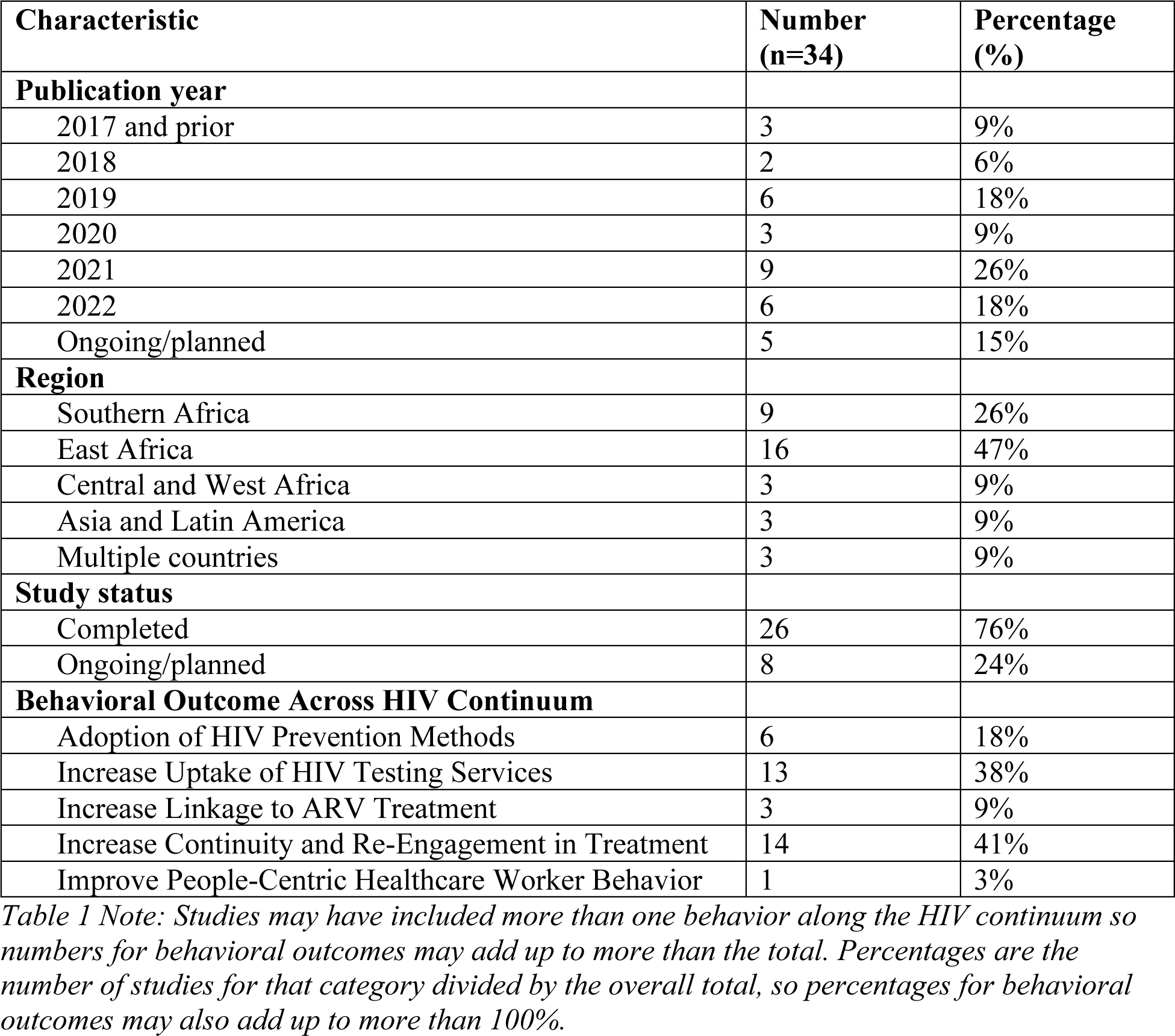
General characteristics of included studies (n=34)

The majority of tested nudges fell into the Attractive (47%) category. The Easy and Social categories each included 13 nudges (38%) while the Timely category included 11 studies (32%) (Table 2).

**Table 2:** Nudge types and mapping studies to EAST Framework (n=34)

|  | **Number**<br>**(n=34)** | **Percentage**<br>**(%)** |
| --- | --- | --- |
| Easy | 13 | 38% |
| Attractive | 16 | 47% |
| Social | 13 | 38% |
| Timely | 11 | 32% |

Within each of the EAST categories, some nudge types were more common. For example, of the 13 studies that had Easy nudges, 12 of them involved simplification, which is 35% of all studies (Table 3).

**Table 3:** Nudge types and mapping studies to EAST Framework (n=34)

|  | **Number**<br>**(n=34)** | **Percentage**<br>**(%)** |
| --- | --- | --- |
| **Easy** | **13** | **38%** |
| Simplification | 12 | 35% |
| Defaults | 1 | 3% |
| **Attractive** | **16** | **47%** |
| Reframing: gain frame | 7 | 21% |
| Reframing: loss frame | 2 | 6% |
| Priming | 1 | 3% |
| Affect effect | 1 | 3% |
| Goal-gradient effect | 1 | 3% |
| Attractive: other nudges | 7 | 47% |
| **Social** | **13** | **38%** |
| Social proof | 11 | 32% |
| Commitment devices | 2 | 6% |
| **Timely** | **11** | **32%** |
| Plan making | 4 | 12% |
| Reminders | 6 | 18% |
| Salience | 3 | 9% |
| Fresh start effect | 1 | 3% |

Most studies identified have been completed and evaluated but eight studies were identified that are planned or ongoing (Table 4).

**Table 4:** Single Nudge Intervention vs Multiple/Mixed Nudge Intervention (n=34)

|  | Number<br>(n=34) | Percentage (%) |
| --- | --- | --- |
| **Completed Studies** | **26** | **76%** |
| Single | 17 | 50% |
| Mixed | 9 | 26% |
| **Ongoing/planned studies** | **8** | **24%** |
| Single | 2 | 6% |
| Mixed | 6 | 18% |

### Easy

Using the EAST Framework, nudges that fit into the “Easy” category include simplification by reducing the ‘hassle factors’ associated with completing a behavior, changing defaults, and simplifying messaging.^5–17^

#### Simplification

Simplifying or eliminating some of the steps that a person needs to complete reduces the ‘hassle factors’ and annoyances that are associated with doing a behavior.^18^ As discussed in detail below, effective simplification has included modifications to program structures or modifications to a product needed to do a behavior.

Many countries have simplified programs for clients by introducing Differentiated Service Delivery (DSD) such as Multi-Month Dispensing, a set of interventions generally offered to individuals who are in treatment and stable. The specifics and details of DSD programs vary widely by country but have a common aim of simplifying services around the when, where, who, and what of care delivery. ‘When’ might reduce the frequency of facility visits, ‘where’ might shift services to community-based options, ‘who’ might provide a wider range of individuals to provide services, and ‘what’ might modify the support structure or testing schedule, among other possibilities. It is outside of the scope of this review to fully discuss the diversity and nuances of DSD programs, but it is important to acknowledge overall program improvements because of them.^19–27^

Program simplification involves making it easier for clients to use services, including moving services into the community or facilitating access to services. The ‘Girl Champ’ program in Eswatini created empowering and fun events at health clinics for adolescent girls and young women (AGYW). Crucially, they transported AGYW to these events at the clinics and provided optional ways for them to interact with service providers during the event, significantly reducing the hassles of seeking services, and while making AGYW more comfortable returning to the clinics. ‘Girl Champ’, which increased the proportion of AGYW-attended HIV testing and counseling before and the after the events, involved additional nudges including using a gain-frame for messaging (an Attractive nudge). Images and branding featured a lioness and the phrase “Fierce, not fearful”.^12^

Service simplification that was possible because of product improvements included self-test kits that an individual can use at home, which reduce the hassles factors and discomfort associated with going to a clinic. Sending notifications on the availability of and providing self-test kits increased testing rates for both male truckers^6^ and female sex workers in Kenya.^7^ When pregnant women were provided with self-test kits to give to their partners, partners were more likely to test in Kenya.^8^ In Malawi, partners who received self-test kits coupled with phone reminders were also more likely to test.^9^

Additional product simplification has included modifying the delivery mechanism of PrEP from a daily pill to either a monthly injection or monthly vaginal ring. Improvements in adherence largely drove the 92% improvement in effectiveness of injections compared to daily pills in a multi-country trial.^10^ In South Africa and Zimbabwe a study from 2019-2021 demonstrated that the monthly vaginal ring had higher moderate adherence by AGYW (77.8%) after 26 months compared to daily pills (58.6%).^11^

#### Defaults

Defaults are pre-set choices that assign an individual to an outcome unless they actively choose another option. They do not limit choice and are sometimes referred to as opt-in or opt-out choices.^18,28^ Effective default nudges have included automatically enrolling individuals in care or automatically providing them with a product.

An example of a default nudge is Universal Test and Treat, a policy that automatically enrolls an individual who tests positive for HIV onto ART and into a care and treatment program regardless of CD4 count or WHO clinical stage. While it was implemented for clinical reasons because it improved outcomes for patients, the policy also fits into the category of Easy nudges. Universal Test and Treat simplified the next steps for a person who tested positive for HIV and improved linkage to care.^29^ So even though it was not implemented because it was a nudge, it is important to acknowledge the improvements it provided to HIV programs and clients.

An additional example of a default nudge that moved the needle on HIV prevention was providing AGYW with HIV self-test kits by default as part of purchases for sexual and reproductive health products. In addition to a default nudge, the “Malkia Klabu” (Queen Club) project used human-centered design to identify and target AGYW’s motivations (an Attractive nudge) to brand sexual and reproductive health products and put them in accredited drug dispensing outlets (ADDOs). Over the course of the intervention, significantly more HIV self-test kits were distributed to AGYW in the ADDOs with the products compared to the AGYW in the control group who, despite the self-test kits also being free to them, had to overcome any discomfort and actively request one.^12,30^

### Attractive

The “Attractive” category of nudges includes framing messaging in various ways that focus attention on those messages or in a way that targets an individual’s motivations and aspirations. It also includes appropriately designed rewards or sanctions.^5,12–14,31–40^

#### Reframing: gain-frame

Focusing on the positive outcomes, for example, highlighting benefits or a positive future, is known as a gain-frame.^41,42^ Gain-frame nudges have focused on how happy or unchanged an individual’s life would be when they are virally suppressed.

Undetectable=Untransmittable (U=U) messaging has been used in HIV programs to improve retention in care, but it has also recently been used to increase testing.^28^ In a recent study in South Africa, called ‘iMpilo’, men who received gain-frame U=U messaging were 1.89 times more likely to get tested (22% compared to 13%) than those who received standard of care messaging promoting testing.^31^ The specific messaging was co-designed with men to target their fears and motivations and included phrasing such as, “It [ART] protects you even if you don’t use a condom. Even if you’re drinking”.^43^ Gain-frame U=U messaging focused on being able to have a fulfilling life on ART has also had positive effects with men and other populations around testing in Nigeria^32, 45^ and Tanzania.^33^

#### Reframing: loss-frame

Whereas gain-frame messaging focuses on the positive future, loss frame messaging takes a different approach and highlights what someone would lose if they do not do a specific action. Gain-frame and loss-frame messaging target different behavioral barriers. Gain-frame messaging like U=U typically targets the paralyzing fears (called ostriching) that people have concerning HIV^34, 44^ by highlighting that they are not going to come true. Loss-frame messaging comes from people displaying loss aversion while making decisions – that losing something of value they already own or already feel they have is more motivating than potentially gaining something of equal value.^30, 41^

A study in Uganda compared a gain-frame and loss-frame approach using small non-monetary incentives as the potential gain or loss. They found that there were not substantial differences in testing rates between individuals who received either the gain or loss frame.^35^ A planned study in South Africa intends to test loss-aversion language to improve retention in care and to improve re-engagement in care for adults who have missed an appointment.^38^ The study is in the planning stages and the details of the loss aversion messaging have not yet been released.

#### Targeted messaging and branding

Creating targeted messaging and branding, frequently using human-centered design to understand motivations and aspirations and to co-create content, may increase the likelihood that individuals notice, identify with, and potentially act on the communication they receive.

The Mpilo project in South Africa applied human-centered design to understand men’s fears and motivations and to design a program to improve retention in care. Football (soccer) metaphors were heavily used in the program to target men’s motivations and interests, such as “I can help you get back in the game.” Men were branded as ‘coaches’ and they provided mentoring and support to ‘teams of players’ by reaching out to newly diagnosed men in their community and to men identified as being lost to follow-up by clinics. In addition to the program being an Attractive nudge, it included social proof, a Social nudge (discussed further below), since the ‘coaches’ themselves were living with HIV and were positively modeling their lives. During the observational pilot, the short term retention rate for men in the program was 80%, approximately 20 percentage points higher than the average rate in the community.^34^

Tailoring images and branding was also an element of multiple nudge interventions that increased positive behaviors in AGYW. Co-design was used in the ‘Malika Kubla’ pilot in Tanzania that increased AGYW purchases of sexual and reproductive health products^12^ and in the ‘Girl Champ’ program in Eswatini that increased the proportion of AGYW who attended HIV testing and counseling.^5^

#### Other Attractive Nudges

An additional Attractive nudge included allowing youth to set their own goals for medication adherence in Uganda, which increased the likelihood they met those targets.^36,46^ Attractive nudges that are currently being planned and/or evaluated are gamification in phone-based apps targeting youth in Nigeria^39^ and Ghana^40^ as well as messaging to adults in South Africa highlighting altruism to improve continuity of treatment and re-engagement in care.^38^

### Social

Nudges that fit into the “Social” category mainly include different types of social proof.^8, 9, 13, 34, 38–40, 47–52^

#### Social Proof

Being told what behavior others, particularly those in key reference groups, are doing or support doing provides cues for individuals that they frequently use to adjust their own behavior.^53^ Social proof can mean giving people information that the majority of people in the reference group do or support doing, or it could include showing people that those individuals are doing the behavior.

A quasi-experimental study in Tanzania used a growing Baobab tree to visually show that people in the care and treatment program were meeting their adherence targets, a form of social proof for patients in the program.^13^ When an individual attended their third on-time visit they received a sticker (a ‘leaf’) with empowering words such as ‘brave’ or ‘courageous’ on it. They could put the ‘leaf’ on the Baobab tree, a symbol of health and strength for the community. Individuals attending their clinic visits saw the tree growing with each leaf. The tree transformed the hidden behavior of sticking to HIV appointments into a visual behavior for patients. After six months, individuals who attended the clinic in the two weeks per month when the intervention was visible (i.e., the exposed) were significantly more likely to be in care compared to individuals who attended the clinics in the non-intervention weeks and did not see the tree or have the ability to earn stickers/leaves (i.e., the non-exposed). The attendance rate for the exposed group was 85% compared to 79% for the non-exposed group. Individuals in the exposed group were also more likely to achieve Medication Possession Ratio >95% (MPR>95%), and they were more likely to believe that other patients were adhering to treatment.

Another form of social proof that has been tested in HIV interventions is telling people about others’ behavior to change the perceived social norm. Using cross-sectional data, two studies in Uganda found a significant correlation between adults’ perception of others’ behavior and their own behavior. Men who believed that HIV testing was not the social norm were 2.6 times more likely to have never been tested for HIV compared to men who believed that testing was the social norm.^37^ Adults on treatment who believed that suboptimal adherence was the social norm (as measured by the number of pills taken in the previous week) were less likely to report optimal adherence themselves compared to adults who believed optimal adherence was the local norm.^38^

### Timely

The “Timely” category of nudges includes different degrees of plan making as well as sending reminders to individuals.^9,12, 16, 17, 40, 51, 54–58^

#### Plan making

Making a plan or determining the course of action you will need to do to achieve a goal solidifies the intention and may include setting up or visualizing the steps needed to complete the action. A concrete plan might include identifying the steps you will need to do and formalizing at least one of the steps, for example by making an appointment. A concrete plan can also include determining alternatives if you deviate from any of the original steps, whereas a weaker plan might be privately deciding to do something.^18, 59^

Appointments, a plan that you will be somewhere at a specific date and time, can be classified as a concrete plan. (They can also be considered a type of commitment device – a Social nudge – since you are making a commitment to someone else^60^). A clinical trial in urban Malawi randomized sober men at bars and nightclubs to, among other options, the offer to schedule an appointment for an HIV test that included a phone reminder two days before the appointment.

Overall, 27% of men in the arm that offered appointments got tested, significantly higher than the rate of 11% in the control group that was provided standard of care testing information. Looking closer at the group that was offered appointments, two-thirds of the men in the appointment offer arm decided to schedule an appointment, and 34% of those who made an appointment got tested.^51^ A study in Thailand had similar results. Adults at a clinic who tested negative were randomized into one of three arms for a subsequent HIV test: making an appointment before they left the clinic, being sent a reminder to make an appointment, and a standard of care arm. The testing rate was highest in the appointment arm, with 37% getting tested, compared with 11% in the standard of care arm and 19% in the reminder arm.^54^

A small study in Uganda investigated a less concrete type of plan making by providing men with a calendar of community health event dates. Both arms received the calendar and one arm included a plan making prompt that suggested they circle the date of the specific event they would attend for HIV testing and select a morning or afternoon visit. The study was underpowered because of the limited number of men available in the communities, and the difference in testing rates of 77.1% in the plan making arm compared to 74.9% in the control arm was not significantly significant. Of note was that the sub-group who received the calendar and prompt less than 41 days before the next health event had a testing rate of 80.7%.^55^

#### Reminders

Reminders cut through the noise and natural forgetfulness of busy lives and provide a prompt for a desired or preplanned action.^61^

Some of the studies included reminders as part of the overall intervention, but few looked at only reminders. For example, the study in Malawi that provided HIV self-kits for partners of pregnant women included reminders to take the test^9^. The studies that evaluated scheduled appointments for HIV tests also included reminders.^51, 54^

### Limitations

This scoping review focused on a subset of HIV behaviors and populations that have recently received renewed focus because they have been identified as not reaching targets or have decelerating progress. While this limits the number of studies included, this targeted focus allows for a deeper discussion of the studies and their impact on these crucial populations.

Additionally, this scoping review excluded studies whose primary or sole intervention was financial or non-monetary nudges, e.g., prizes in place of financial incentives. Because of limited resources and lack of sustainability, financial nudges are viewed as either not feasible, difficult to scale, or outside the range of program options by policy makers and program managers, the primary audience for this scoping review. Therefore financial nudges were excluded from this scoping review, but they are included in Andrawis et al.^62^

Additional limitations are based on the structure of the literature itself. Many studies focused on broad populations, such as all adults, and did not conduct sub-analyses on men and women or older adults and younger adults. This lack of differentiation between populations means it is unclear if or how a target population responded to an intervention, adding more uncertainty to future implementation.

Additionally, articles were authored by scientists from the variety of fields that behavioral economics encompasses: economists, psychologists, sociologists, anthropologists, social and behavior change specialists, and decision scientists. This diversity meant that terms and study reporting varied widely. To be inclusive of all potential nudges, EL reviewed studies that may not have used standard BE terminology but after examining the details of the intervention, classified them into a specific type of nudge and into the EAST Framework. Many of the studies did not report a detailed behavioral diagnosis, i.e., explaining the factors that were making it difficult for individuals to do the behavior, and of the few studies that did, even fewer reported their diagnosis using BE terminology. Therefore this scoping review has not included or summarized common diagnoses for each behavior.

## DISCUSSION AND IMPLICATIONS

This scoping review shows that there are significant gaps in the applications of nudges along the HIV continuum. There are nudges that have been shown to be effective but have not yet been implemented at a larger scale, a significant missed opportunity for HIV epidemic control.

It is important to discuss the effect sizes of many of the statistically significant nudges in the studies. Nudges typically have small effect sizes but are almost always low-cost, meaning that it is relatively easy to try to implement and test them. The nudges tested in these studies tend to have relatively modest effects, but even small changes can potentially move the needle for behaviors that have persistently been lagging. Moving the needle on these behaviors can have multiplier effects that help reduce the overall spread of HIV.

There is a range of planned or ongoing but not yet evaluated nudges that the scoping review was able to identify that, when completed, will fill some of the current knowledge gaps of nudges and HIV programs. These nudges include testing the fresh start effect (a Timely nudge) that uses a key date, such as the first of the month or the start of a season, to give someone a mental reset which can motivate them to change their behavior, especially around re-engagement in care.^58, 63, 64^ Gamification and game-based text messaging is also being tested in various contexts, particularly with youth,^39, 40^ to increase engagement in care and reduce interruptions in treatment. Gamification interventions tend to rely heavily on non-monetary rewards like “badges” and being able to advance to a higher level in an online or phone-based app, intermittent recognition, and they frequently use social comparison tools such as leaderboards.

There is a striking lack of studies that apply nudges to improve client-centric care or change provider behavior in LMICs. However, there are strong hints as to a path forward from active studies in LMICs applying nudges to non-HIV contexts as well as significant results from non-LMIC contexts. A behaviorally informed intervention to improve respectful maternity care is being evaluated in Zambia that includes reframing (Attractive), plan making (Timely), and goal-gradient nudges (Attractive).^65^ A Cochrane review of nudges on provider behavior highlighted that the majority of included studies had statistically significant effects on provider behavior.^66^ Effective interventions included action alerts and decision supports for doctors, simplifying decision-making and making it salient, which increased guideline-concordant care.^67–70^

## CONCLUSION

There is strong evidence that the incorporation of nudges into HIV programs can improve behavior along the HIV continuum. However, behavioral economics is not a silver bullet. While not all studies significantly changed behaviors, those that did help form a pool of potentially effective interventions that have not been sufficiently harnessed in HIV programs. This set of interventions should not be seen as replacements for current programs but rather as innovative ways to enhance programs.

The effect sizes of nudges may be modest, but their low cost and relative ease of implementation mean that these small gains can help to tackle the last mile challenges. The HIV community needs to continue designing and incorporating nudges into programs while evaluating their impacts using implementation science and other rigorous evaluation methods. Incorporating nudges into programs now can help move the needle to and beyond the 95-95-95 targets and end the epidemic as a public health threat by 2030.

## Data Availability

All data produce in the present work are contained in the manuscript.

## Supplement 1: List of search terms

The keyword “HIV” plus one of the following keywords were searched:

behavioral economics
behavioral design
behavioral insights
behavioral bias
cognitive bias
nudge
commitment devices
precommitment
reminders
loss aversion
present bias
planning
appointments
implementation intentions
heuristics
mental shortcut
mental model
framing
priming
inertia
friction
hassle factors
incentives
recognition
defaults
status quo
gamification
social proof
scarcity
optimism bias
appraisal bias
availability bias
empathy gap
licensing
ego depletion
salience
fresh start effect
endowment effect
health worker behavior
respectful care

## Supplement 2: Nudge details and EAST Classification by study

**Table S2:** Nudge details and EAST Classification by study.

| Citation | Reference and Title | Status | Easy | Attractive | Social | Timely |
| --- | --- | --- | --- | --- | --- | --- |
| 5 | Project Last Mile and the development of the Girl Champ brand in eSwatini: engaging the private sector to promote uptake of health services among adolescent girls and young women | Completed | simplification | gain-frame; targeted branding |  |  |
| 6 | Announcing the availability of oral HIV self-test kits via text message to increase HIV testing among hard-to-reach truckers in Kenya: a randomized controlled trial | Completed | simplification |  |  |  |
| 7 | A randomized controlled trial to increase HIV testing demand among female sex workers in Kenya through announcing the availability of HIV self-testing via text message | Completed | simplification |  |  |  |
| 8 | Promoting partner testing and couples testing through secondary distribution of HIV self-tests: a randomized clinical trial | Completed | simplification |  | social proof |  |
| 9 | HIV self-testing alone or with additional interventions, including financial incentives, and linkage to care or prevention among male partners of antenatal care clinic attendees in Malawi: an adaptive multi-arm, multi-stage cluster randomised trial | Completed | simplification |  | social proof | phone reminders |
| 10 | Characterization of human immunodeficiency virus (HIV) infections in women who received injectable cabotegravir or tenofovir disoproxil fumarate/emtricitabine for HIV prevention: HPTN 084 | Completed | simplification |  |  |  |
| 11 | Adherence to the dapivirine vaginal ring and oral PrEP among adolescent girls and young women in Africa: interim results from the REACH study | Completed | simplification |  |  |  |
| 12 | AmbassADDOrs for Health and Malika Kubla | Completed | simplification | affect effect; targeted branding |  | salience |
| 13 | Pilot study of a multi-pronged intervention using social norms and priming to improve adherence to antiretroviral therapy and retention in care among adults living with HIV in Tanzania | Completed | simplification | priming | social proof |  |
| 14 | PrEP My Way | Ongoing/<br>Planned | simplification | targeted branding |  |  |
| 15 | Evaluating the Acceptability and Feasibility of the B-ok bead bottles in Kwazulu-Natal | Ongoing/<br>Planned | simplification |  |  |  |
| 16 | Febrisol Stickers | Ongoing/<br>Planned | simplification |  |  | salience |
| 17 | Prioritizing Retention Efforts Using Data Intelligence and Cohort Targeting in South Africa | Ongoing/<br>Planned | simplification |  |  | plan making; salience |
| 31 | Undetectable=untransmittable (U=U) messaging increases uptake of HIV testing among men: results from a pilot cluster randomized trial | Completed |  | gain-frame; targeted branding |  |  |
| 32 | Reaching Impact, Saturation, and Epidemic Control (RISE) | Completed |  | gain-frame |  |  |
| 33 | Furaha Yangu (My Happiness) | Completed |  | gain-frame |  |  |
| 34 | Uptake and Short-Term Retention in HIV Treatment Among Men in South Africa: The Coach Mpilo Pilot Project | Completed |  | gain-frame; targeted branding | social proof |  |
| 35 | Comparative effectiveness of novel non-monetary incentives to promote HIV testing: a randomized trial | Completed |  | gain-frame; loss-frame |  |  |
| 36 | Moving the Goalpost Closer: Do Flexible Targets Improve the Behavioral Impact of Incentives? | Completed |  | goal-gradient |  |  |
| 37 | Appraising praise: experimental evidence on positive framing and demand for health services | Completed |  | gain-frame |  |  |
| 38 | Behavioural Text Messaging to Improve Retention in Care for Patients on Antiretroviral Therapy in Gauteng province in South Africa | Ongoing/<br>Planned |  | loss-frame;<br>altruism | social proof |  |
| 39 | PEERNaija: A Gamified mHealth Behavioral Intervention to Improve Adherence to Antiretroviral Treatment Among Adolescents and Young Adults in Nigeria | Ongoing/<br>Planned |  | gamification | social proof |  |
| 40 | Preferences for a Game-Based SMS Adherence Intervention Among Young People Living with HIV in Ghana: A Qualitative Study | Ongoing/<br>Planned |  | gamification | social proof | reminders |
| 47 | Actual versus perceived HIV testing norms, and personal HIV testing uptake: a cross-sectional, population-based study in rural Uganda | Completed |  |  | social proof |  |
| 48 | Perceptions About Local ART Adherence Norms and Personal Adherence Behavior Among Adults Living with HIV in Rural Uganda | Completed |  |  | social proof |  |
| 49 | A randomized controlled trial study of the acceptability, feasibility, and preliminary impact of SITA (SMS as an Incentive To Adhere): a mobile technology-based intervention informed by behavioral economics to improve ART adherence among youth in Uganda | Completed |  |  | social proof |  |
| 50 | Behavioral intention to initiate antiretroviral therapy (art) among Chinese HIV-infected men who have sex with men having high CD4 count in the era of “Treatment for All” | Completed |  |  | social proof |  |
| 51 | Appointments: A more effective commitment device for health behaviors | Completed |  |  | commitment device | plan making; reminders |
| 52 | Using Incentives and Nudging to Improve Non-Targeted HIV Testing in Ecuador: A Randomized Trial | Completed |  |  | commitment device |  |
| 54 | Appointment reminders to increase uptake of HIV retesting by at-risk individuals: a randomized controlled study in Thailand | Completed |  |  |  | plan making; reminders |
| 55 | Planning prompts to promote uptake of HIV services among men: a randomised trial in rural Uganda | Completed |  |  |  | plan making |
| 56 | Effect of SMS reminders on PrEP adherence in young Kenyan women (MPYA study): a randomised controlled trial | Completed |  |  |  | reminders |
| 57 | Text messaging for improving antiretroviral therapy adherence: no effects after 1 year in a randomized controlled trial among adolescents and young adults | Completed |  |  |  | reminders |
| 58 | “Fresh start” text messaging to motivate recipients of care with treatment interruptions to re-initiate antiretroviral therapy in the Capricorn District, South Africa | Ongoing/<br>Planned |  |  |  | fresh start |

